# Development of a Novel Risk Score for Predicting One-Year Mortality Risk in Patients with Atrial Fibrillation using XGBoost-Assisted Feature Selection

**DOI:** 10.1101/2024.01.09.24301080

**Authors:** Bin Wang, Feifei Jin, Han Cao, Qing Li, Ping Zhang

**Affiliations:** Department of Cardiology, Beijing Tsinghua Changgung Hospital, School of Clinical Medicine, Tsinghua University, Beijing, China; Trauma Medicine Center, Peking University People’s Hospital, Beijing, China; Key Laboratory of Trauma treatment and Neural Regeneration, Peking University, Ministry of Education, Beijing, China; National Center for Trauma Medicine of China, Beijing, China; Medical Data Science Center, Beijing Tsinghua Changgung Hospital, School of Clinical Medicine, Tsinghua University, Beijing, China

**Keywords:** Atrial fibrillation, Mortality, Risk score, Machine learning, Risk factors

## Abstract

**Background:** There is a lack of tools specifically designed to assess mortality risk in patients with atrial fibrillation (AF). The aim of this study was to utilize machine learning methods for identifying pertinent variables and developing an easily applicable prognostic score to predict 1-year mortality in AF patients.

**Methods:** This single-center retrospective cohort study based on the Medical Information Mart for Intensive Care-IV (MIMIC-IV) database focused on patients aged 18 years and older with AF. The study thoroughly scrutinized patient data to identify and analyze variables, encompassing demographic variables, comorbidities, scores, vital signs, laboratory test results, and medication usage. The variable importance from XGBoost guided the development of a logistic model, forming the basis for an AF scoring model. Decision curve analysis was used to compare the AF score with other scores. Python and R software were used for data analysis.

**Results:** A cohort of 59,595 AF patients was obtained from the MIMIC-IV database; these patients were predominantly elderly (median age 77.3 years) and male (55.6%). The XGBoost model effectively predicted 1-year mortality (Area under the curve (AUC): 0.833; 95% confidence intervals: 0.826-0.839), underscoring the significance of the Charlson Comorbidity Index (CCI) and the presence of metastatic solid tumors.

The CRAMB score (Charlson comorbidity index, readmission, age, metastatic solid tumor, and blood urea nitrogen maximum) outperformed the CCI and CHA2DS2-VASc scores, demonstrating superior predictive value for 1-year mortality. In the test set, the area under the ROC curve (AUC) for the CRAMB score was 0.756 (95% confidence intervals: 0.748-0.764), surpassing the CCI score of 0.720 (95% confidence intervals: 0.712-0.728) and the CHA2DS2-VASc score of 0.609 (95% confidence intervals: 0.600-0.618). Decision curve analysis revealed that the CRAMB score had a consistently positive effect and greater net benefit across the entire threshold range than did the default strategies and other scoring systems. The calibration plot for the test set indicated that the CRAMB score was well calibrated.

**Conclusions:** This study’s primary contribution is the establishment of a benchmark for utilizing machine learning models in construction of a score for mortality prediction in AF. The CRAMB score was developed by leveraging a large-sample population dataset and employing XGBoost models for predictor screening. The simplicity of the CRAMB score makes it user friendly, allowing for coverage of a broader and more heterogeneous AF population.

## Introduction

Atrial fibrillation (AF) is a prevalent cardiac arrhythmia linked to considerable morbidity and mortality. It is characterized by an irregular and often rapid heart rate, resulting in compromised blood flow and potential complications such as stroke, heart failure, and other cardiovascular events [1]. AF has a broad impact on cardiac function, functional status, and quality of life and is also a risk factor for stroke [2]. Additionally, AF is a significant risk factor for stroke and becomes more prevalent with age, affecting more than 2 million individuals in the United States, 14% to 17% of whom are aged 65 years and older [3]. Individuals diagnosed with AF experienced a 3.7 times greater likelihood of mortality from any cause than the general population [4]. AF represents a significant public health issue due to its considerable impact on morbidity and mortality as well as its economic strain on healthcare systems.

The assessment tool for evaluating the risk of stroke in patients with AF, known as the congestive heart failure, hypertension, age, diabetes mellitus, prior stroke or TIA or thromboembolism, vascular disease, age, sex category (CHA2DS2-VASc) score [5], has been associated with cardiovascular events and mortality in diverse patient groups, including those with diabetes [6] and individuals without AF [7]. Nevertheless, tools specifically designed to assess mortality risk in patients with AF are lacking. Although recent studies have introduced new AF risk scores [8, 9], these scores were developed based on data from clinical trials, limiting their applicability to the broader AF population.

Consequently, additional research is necessary to identify potential models for scoring AF risk. The objective of this study was to employ machine learning methods to identify relevant variables and create an easily applicable prognostic score for predicting 1-year mortality in AF patients.

## Methods

### Study design and setting

This was a single-center retrospective cohort study. The data utilized in this research originated from the Medical Information Mart for Intensive Care-IV (MIMIC-IV version 2.2) database [10, 11]. Over the period from 2008 to 2019, the intensive care unit (ICU) at Beth Israel Deaconess Medical Center admitted more than 50,000 critically ill patients, as documented in MIMIC-IV. Approval for the MIMIC-IV database was granted by the Massachusetts Institute of Technology (Cambridge, MA) and Beth Israel Deaconess Medical Center (Boston, MA), with consent obtained for the initial data collection.

### Study population

The study population included patients aged 18 years and older with a discharge diagnosis of AF. Patients with AF were identified by searching International Classification of Diseases diagnostic terminology in the MIMIC-IV database by matching the keyword "atrial fibrillation". The types of queried AF diagnostic terms were manually reviewed to ensure compliance. The exclusion criterion was lack of patient data on survival outcomes.

### Study variables

The variables examined in the research included the characteristics of the study population, complications, various scores (such as Charlson Comorbidity Index and the CHA2DS2-VASc score), vital signs, and an array of laboratory tests (including routine bloodwork, blood biochemistry, coagulation, blood lipids, cardiac markers, etc.). Additionally, the investigation considered use of vasopressors (norepinephrine, epinephrine, phenylephrine, dopamine, dobutamine, vasopressin, and milrinone), antithrombotic agents (heparin, enoxaparin, warfarin, aspirin, clopidogrel, ticagrelor, rivaroxaban, edoxaban, dabigatran etexilate, fondaparinux sodium, prasugrel, and apixaban), and beta blockers (propranolol, metoprolol, bisoprolol, carvedilol, labetalol, atenolol, and nebivolol) and various other data points. For laboratory test items, summary statistics, including minimum and maximum values during hospitalization, were utilized to derive variables. An indicator column for the respective drug was generated based on whether the drug was used during hospitalization.

### Outcome variable

The primary outcome measured was 1-year mortality. Survival time was calculated by utilizing the date of death information available in the MIMIC-IV database restricted to a 1-year timeframe.

### Machine learning model development and validation

The dataset was randomly partitioned into training and test samples at a 3:1 ratio. To prevent model overfitting, tenfold cross-validation and model calibration techniques were applied. To accommodate varying degrees of missing values in dataset variables, the mainstream machine learning model XGBoost was employed due to its ability to handle missing data. The discriminative performance of the models was assessed using the area under the receiver operating characteristic (ROC) curve. Feature scaling was deemed unnecessary before inputting the data into the model. A total of 164 candidate variables were incorporated into the model training process.

Furthermore, a calibration curve was utilized as a graphical representation to evaluate the concordance between the predicted probabilities and observed outcomes in binary classification models. On the calibration curve, the x-axis denotes the mean predicted probability assigned by the model to a specific class, and the y-axis signifies the observed frequency of positive instances. Ideally, a well-calibrated model produces a calibration curve that closely aligns with the diagonal line (y=x), signifying a perfect correspondence between the predicted probabilities and actual outcomes.

### Development of the scoring scale

The XGBoost model assigned importance to predictor variables, and variables with higher importance were selected based on this ranking. These selected variables were subsequently integrated into a logistic model to construct the scoring model. Manual testing was employed to evaluate the impact of introducing or removing variables on the area under the curve (AUC) of the logistic model in the test set. After striking a balance between AUC performance and the increase in model complexity associated with the number of variables included, the chosen variables for the AF scoring model were ultimately determined. A nomogram was used to construct the finalized AF scores. Decision curve analysis (DCA) was employed to assess the clinical utility and net benefit of the AF scoring model, CCI, and CHA2DS2-VASc scores within the test set [12, 13]. DCA quantifies the net benefit of a clinical prediction model at different risk thresholds, avoiding the simplistic assumptions of all patients being at low or high risk. The superior model is identified by the highest net benefit at the chosen threshold. The flow chart of the study is shown in **Figure 1**.

**Figure 1.**
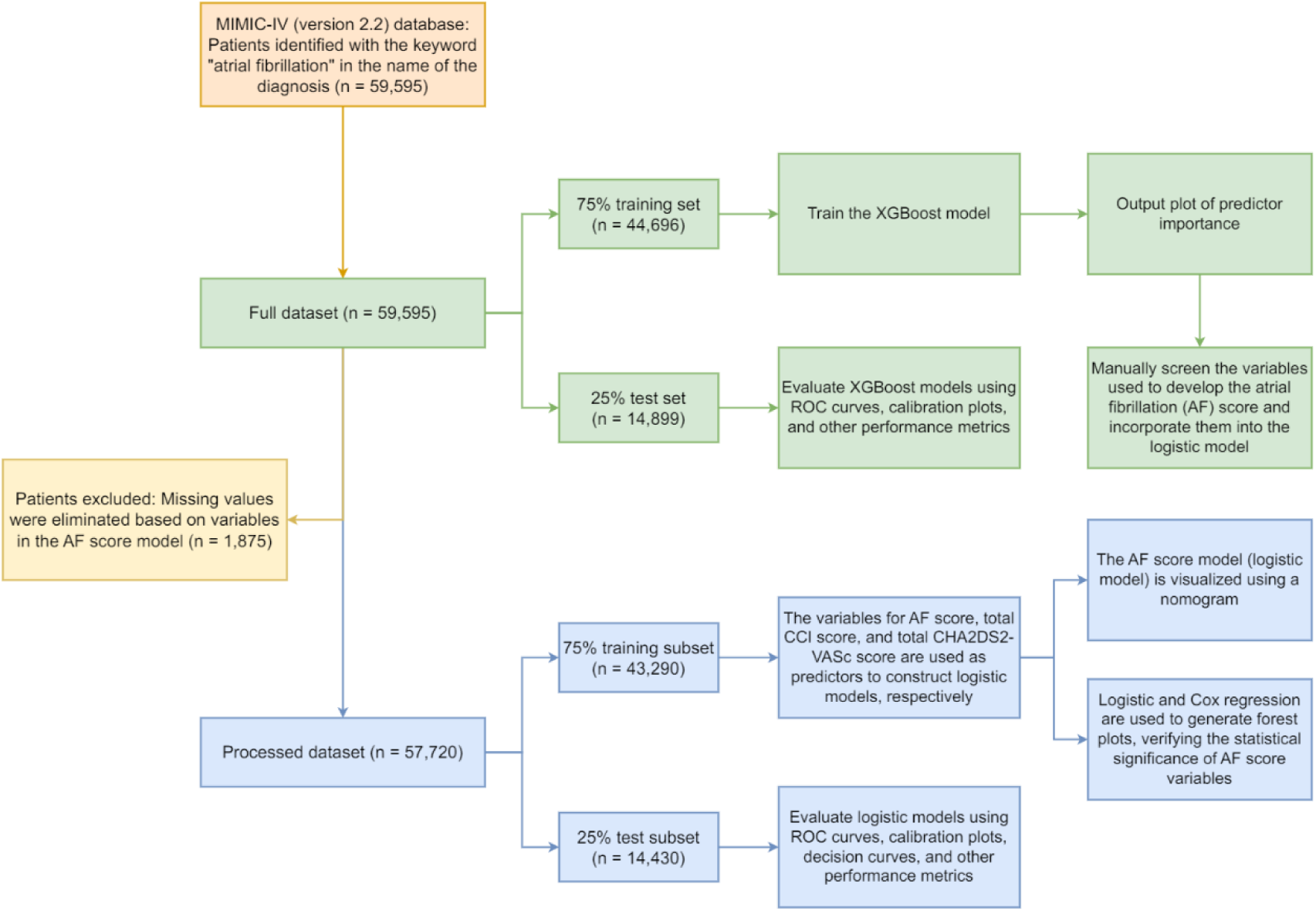
Flow chart of the study. CCI: Charlson comorbidity index; AF: atrial fibrillation

### Data analysis

Python software (version 3.11.5) was used to construct machine learning models, evaluate performance, and generate AUCs and calibration curves. R software (version 4.3.2) was used for logistic and Cox regression analyses, forest plot creation, DCA, and nomogram generation. Baseline characteristics are presented as the mean (standard deviation), median (Q1, Q3), or percentage (%), as determined by the distribution characteristics of the data. The DeLong test was applied to determine whether the AUC of a given prediction significantly differed from that of another prediction [14]. Python was used for descriptive tables [15] and DeLong tests. When constructing the original machine learning model, no handling of missing values was conducted. However, during the development of the logistic model for the AF score, missing values were removed from the dataset based on the variables included in the AF score, as logistic models are unable to manage missing values. In all analyses, statistical significance was defined as a two-sided p value < 0.05.

## Results

### Baseline characteristics

This study enrolled 59,595 individuals diagnosed with AF from the MIMIC-IV database. Among them, 55.6% were male. The cohort had a median age of 77.3 years (with an interquartile range of 68.1-85.3), a median CHA2DS2-VASc score of 4 (interquartile range, 3-5), a median CCI of 6 (interquartile range, 4-8), and a median hospitalization duration of 4 days (interquartile range, 1-7). Additional results are presented in **Table 1**.

**Table1.** Demographic characteristics of the patients in the baseline cohort.

| **Item** | **Category** | **Missing** | **Total (59595)** | **Survivors (33177)** | **Nonsurvivors (26418)** |
| --- | --- | --- | --- | --- | --- |
| **Age, median [Q1, Q3]** | - | 0 | 77.3 [68.1,85.3] | 74.2 [65.3,82.5] | 81.1 [72.3,87.9] |
| **Sex, n (%)** | **Female** | 0 | 26439 (44.4) | 14300 (43.1) | 12139 (45.9) |
|  | **Male** | 0 | 33156 (55.6) | 18877 (56.9) | 14279 (54.1) |
| **In-hospital death, n (%)** | **No** | 0 | 56554 (94.9) | 33177 (100.0) | 23377 (88.5) |
|  | **Yes** | 0 | 3041 (5.1) | - | 3041 (11.5) |
| **Readmission, n (%)** | **No** | 0 | 19728 (33.1) | 12996 (39.2) | 6732 (25.5) |
|  | **Yes** | 0 | 39867 (66.9) | 20181 (60.8) | 19686 (74.5) |
| **Intensive care unit admission, n (%)** | **No** | 0 | 41416 (69.5) | 23724 (71.5) | 17692 (67.0) |
|  | **Yes** | 0 | 18179 (30.5) | 9453 (28.5) | 8726 (33.0) |
| **Hospital length of stay, median [Q1, Q3]** | - | 0 | 4.0 [1.0,7.0] | 3.0 [1.0,6.0] | 4.0 [2.0,8.0] |
| **CHA2DS2-VASc, median [Q1, Q3]** | - | 0 | 4.0 [3.0,5.0] | 3.0 [2.0,5.0] | 4.0 [3.0,5.0] |
| **Charlson comorbidity index, median [Q1, Q3]** | - | 0 | 6.0 [4.0,8.0] | 5.0 [3.0,7.0] | 7.0 [5.0,9.0] |

### Screening variables using the XGBoost model

The XGBoost model showed an AUC and 95% confidence intervals (95% CI) of 0.833 (95% CI, 0.826-0.839) when predicting 1-year mortality in the test set (**Figure 2**). **Figure 3** illustrates the significance of the predictor variables determined by the XGBoost model. Notably, CCI and the presence of metastatic solid tumors were identified as the top two variables, with considerably greater importance than the other variables.

**Figure 2.**
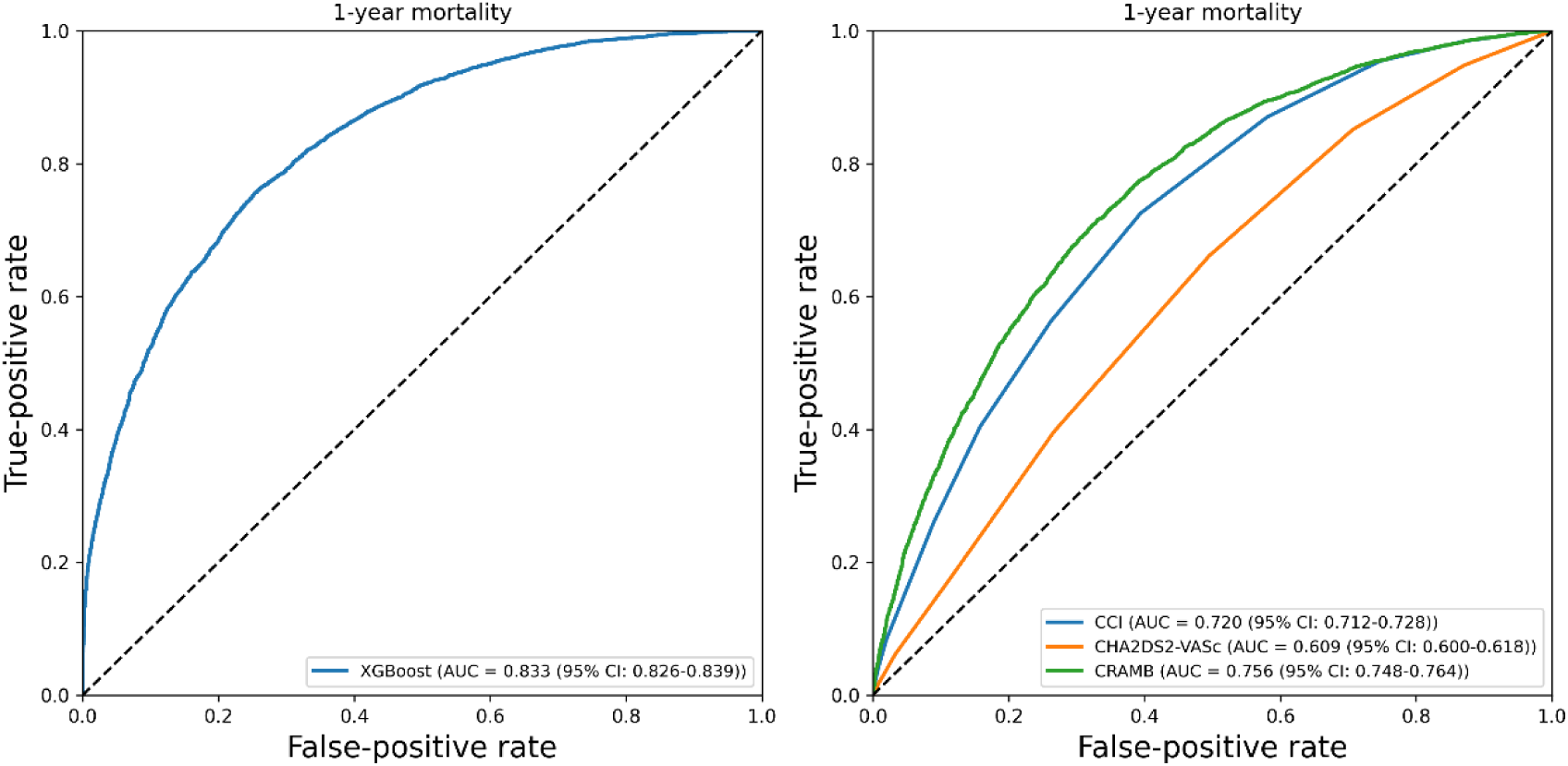
ROC curves for the XGBoost model in the test set and the scoring model in the test set. CCI: Charlson Comorbidity Index; CHA2DS2-VASc score: congestive heart failure, hypertension, age, diabetes mellitus, prior stroke or TIA or thromboembolism, vascular disease, age, sex category; CRAMB score: Charlson comorbidity index, readmission, age, metastatic solid tumor, and blood urea nitrogen maximum.

**Figure 3.**
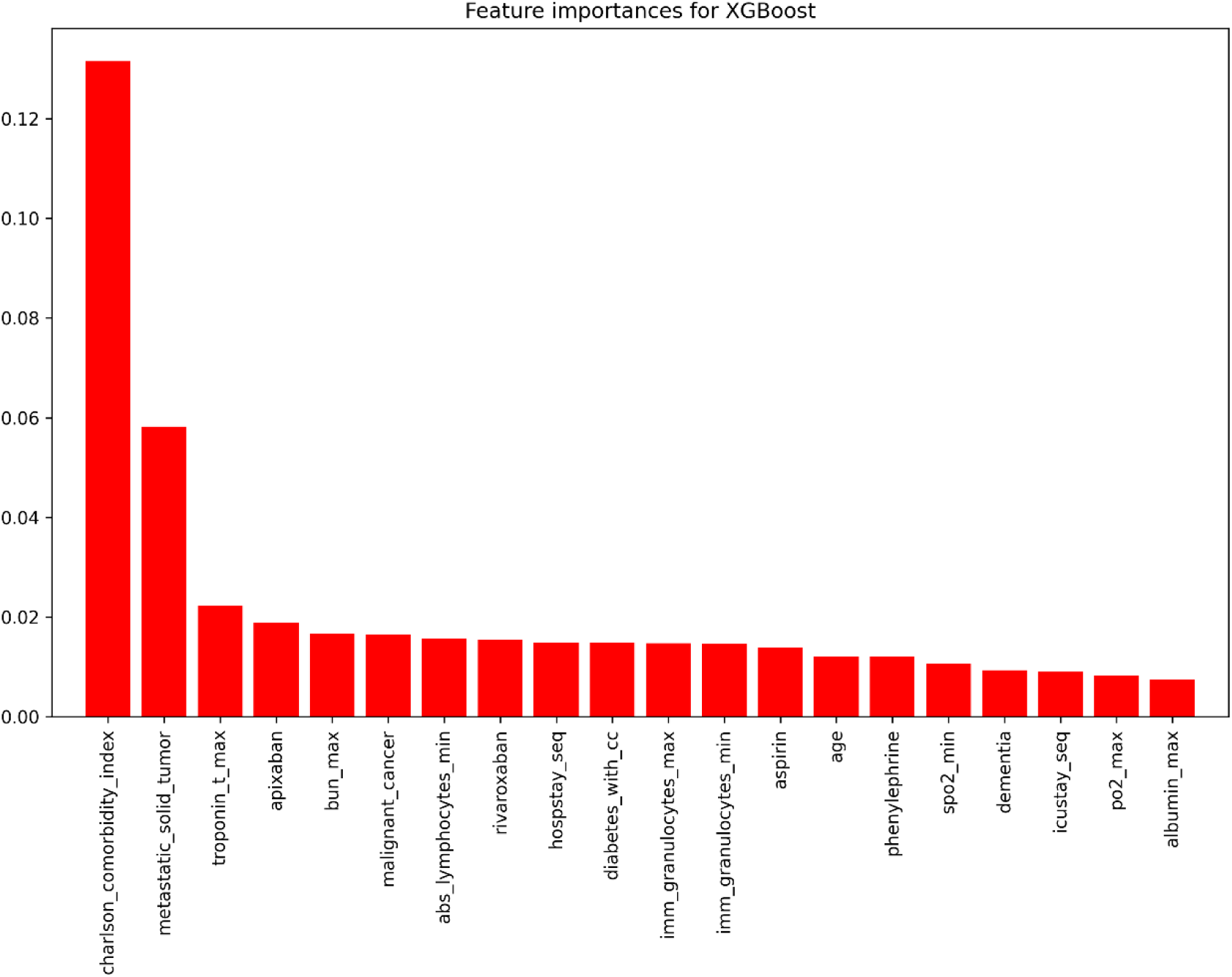
Feature importance values of the XGBoost model in the training set.

### Derivation and evaluation of the AF score

The 1-year mortality risk score for AF was calculated as CRAMB, which represents CCI, readmission, age, metastatic solid tumor, and maximum blood urea nitrogen (BUN) (variable "bun_max") (**Figure 3**). The variable "readmission" was derived from the variable "hospstay_seq" (hospital stay sequence, indicating the number of hospitalizations) for convenient clinical application. Logistic and Cox regression analyses were employed to assess the predictive value of these five variables for the outcome of death, as expressed as odds ratios (ORs). Both the forest plot (**Figure 4**) and the forest plot of hazard ratios (HRs) (**Figure 5**) revealed that these variables were significantly different.

**Figure 4.**
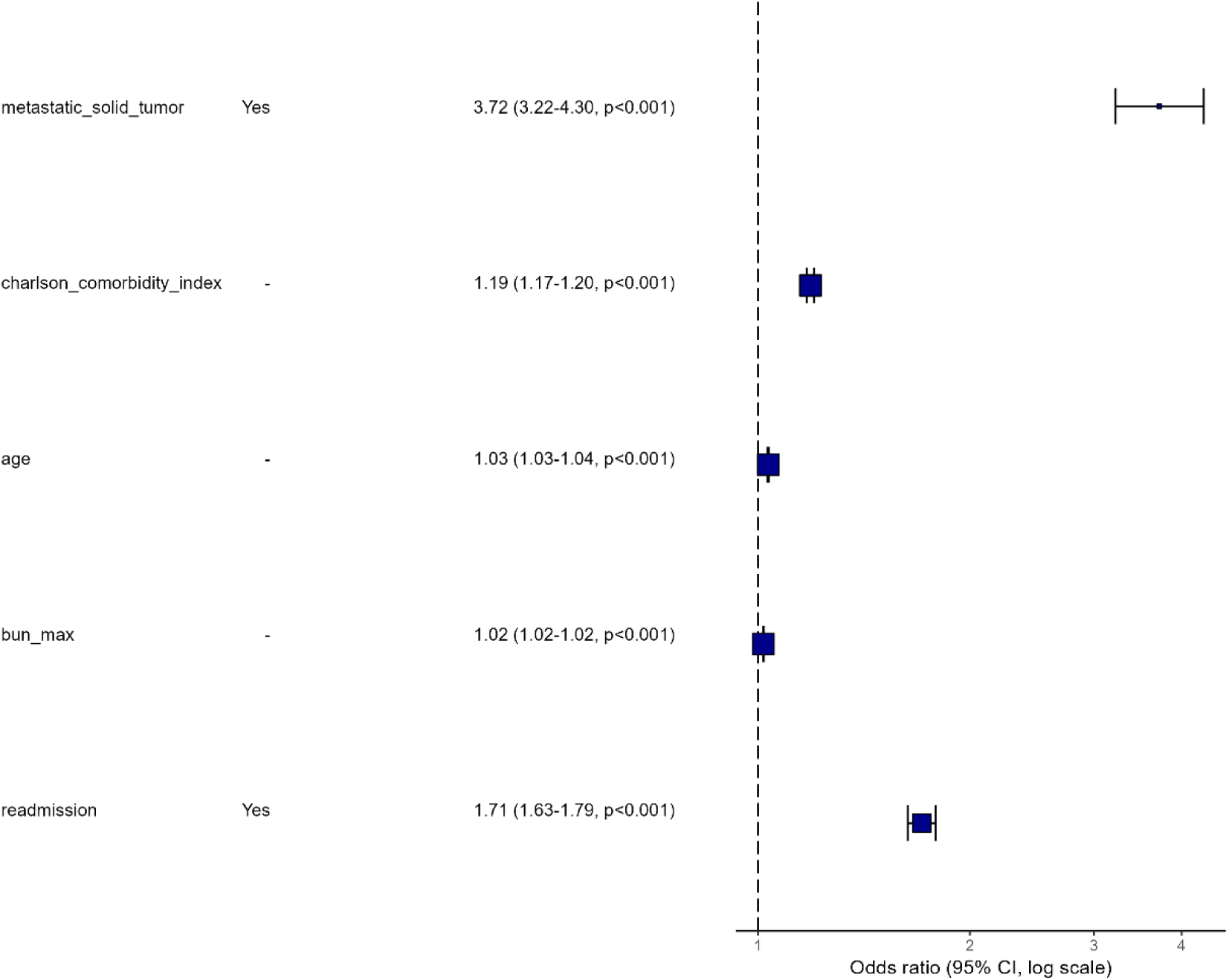
Forest plot of the logistic model (CRAMB score) for predicting 1-year mortality in the training set.

**Figure 5.**
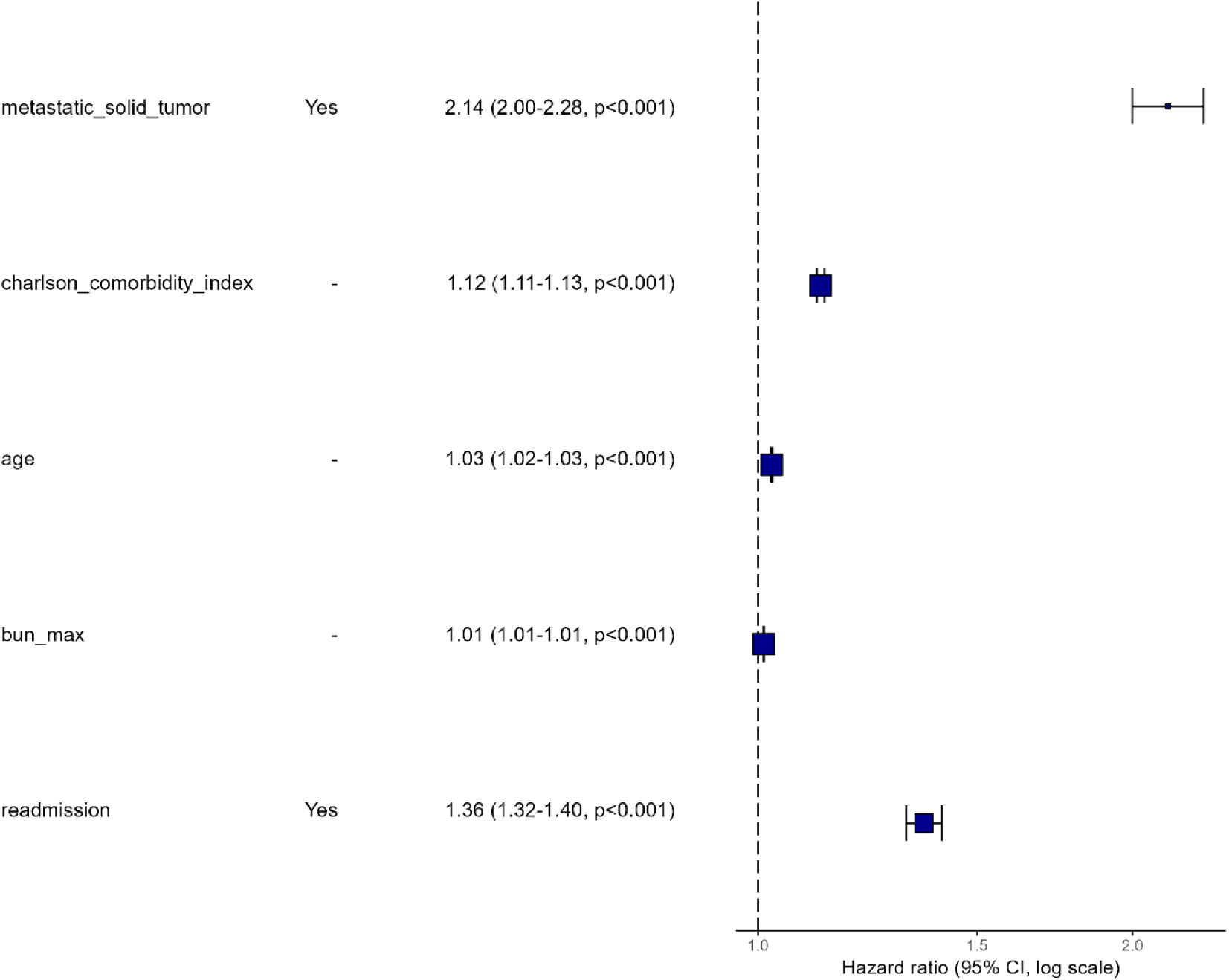
Forest plot illustrating the ability of the Cox regression model to predict 1-year mortality in the training set stratified by the CRAMB score.

A nomogram was used to calculate the CRAMB score (**Figure 6**). In the test set, the area under the curve (AUC) for the CRAMB score was 0.756 (95% CI=0.748-0.764), surpassing CCI, at 0.720 (95% CI=0.712-0.728), and the CHA2DS2-VASc score, at 0.609 (95% CI=0.600-0.618). **Table 2** displays supplementary performance metrics corresponding to these scores. The DeLong test results comparing the CRAMB score with existing scores (CCI and CHA2DS2-VASc) revealed statistically significant differences (p < 0.001), as indicated in **Table 2**. The DCA results provided in **Figure 7** demonstrate that the CRAMB score consistently exhibited a positive and greater net benefit across the entire threshold range than the default strategies, assuming either high or low risk, as indicated by CCI and CHA2DS2-VASc scores, and the hypothesis of not using a scoring system. The calibration plot (**Figure 8**) for the test set indicated that the CRAMB score was well calibrated.

**Figure 6.**
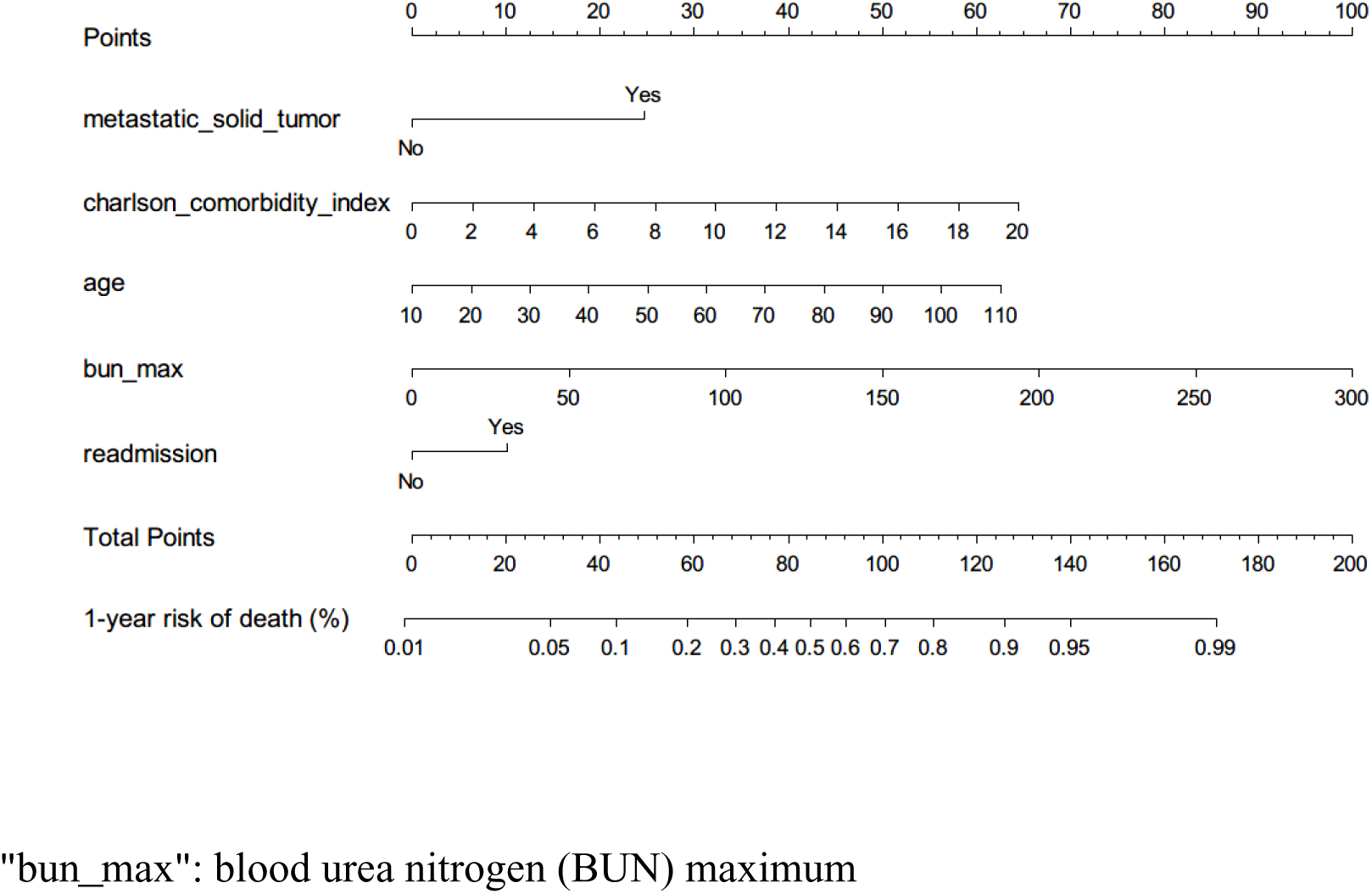
Nomogram of the logistic model (CRAMB score) for predicting 1-year mortality in the training set.

**Figure 7.**
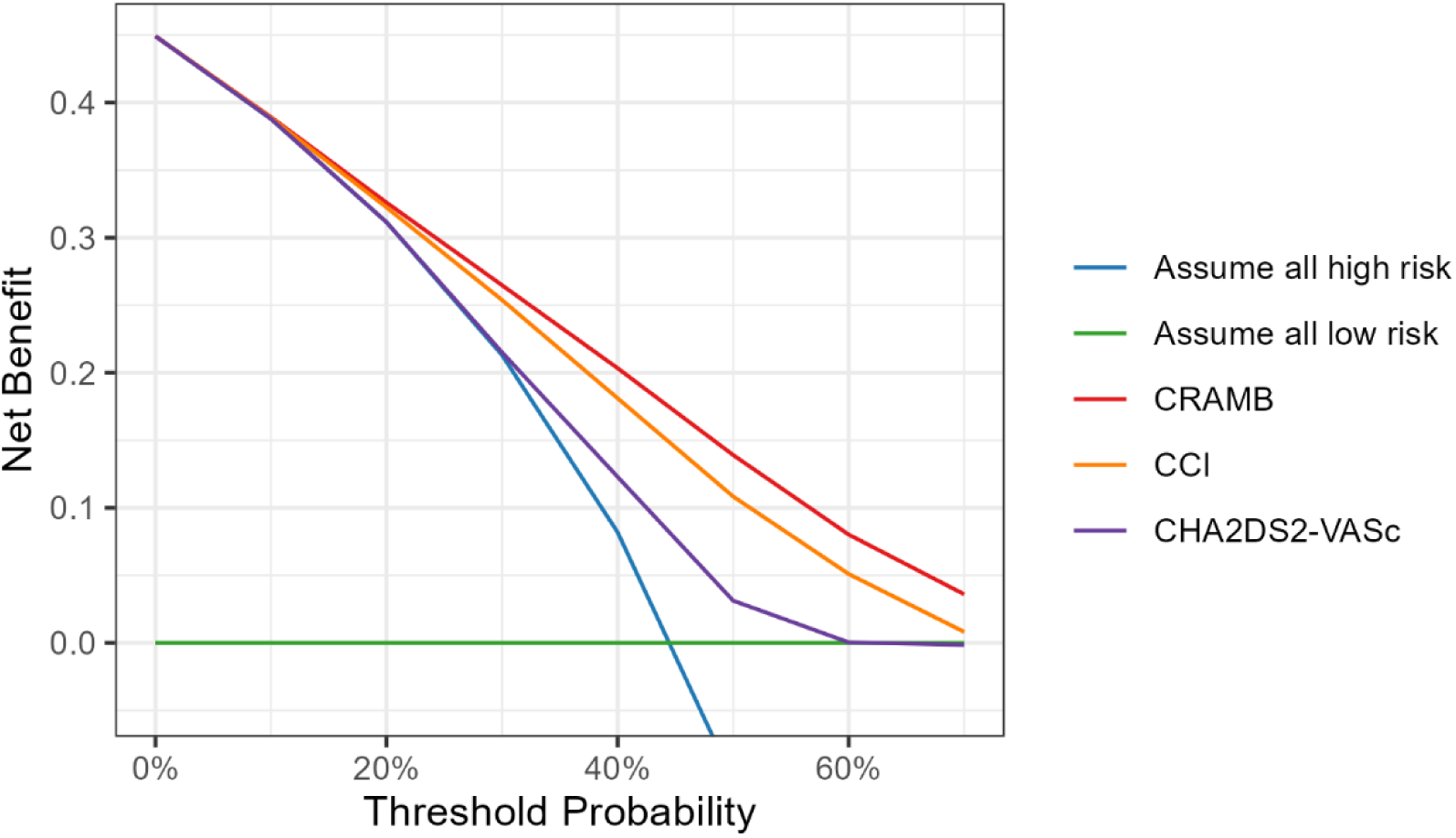
Decision curve analysis of various scores in the training set.

**Figure 8.**
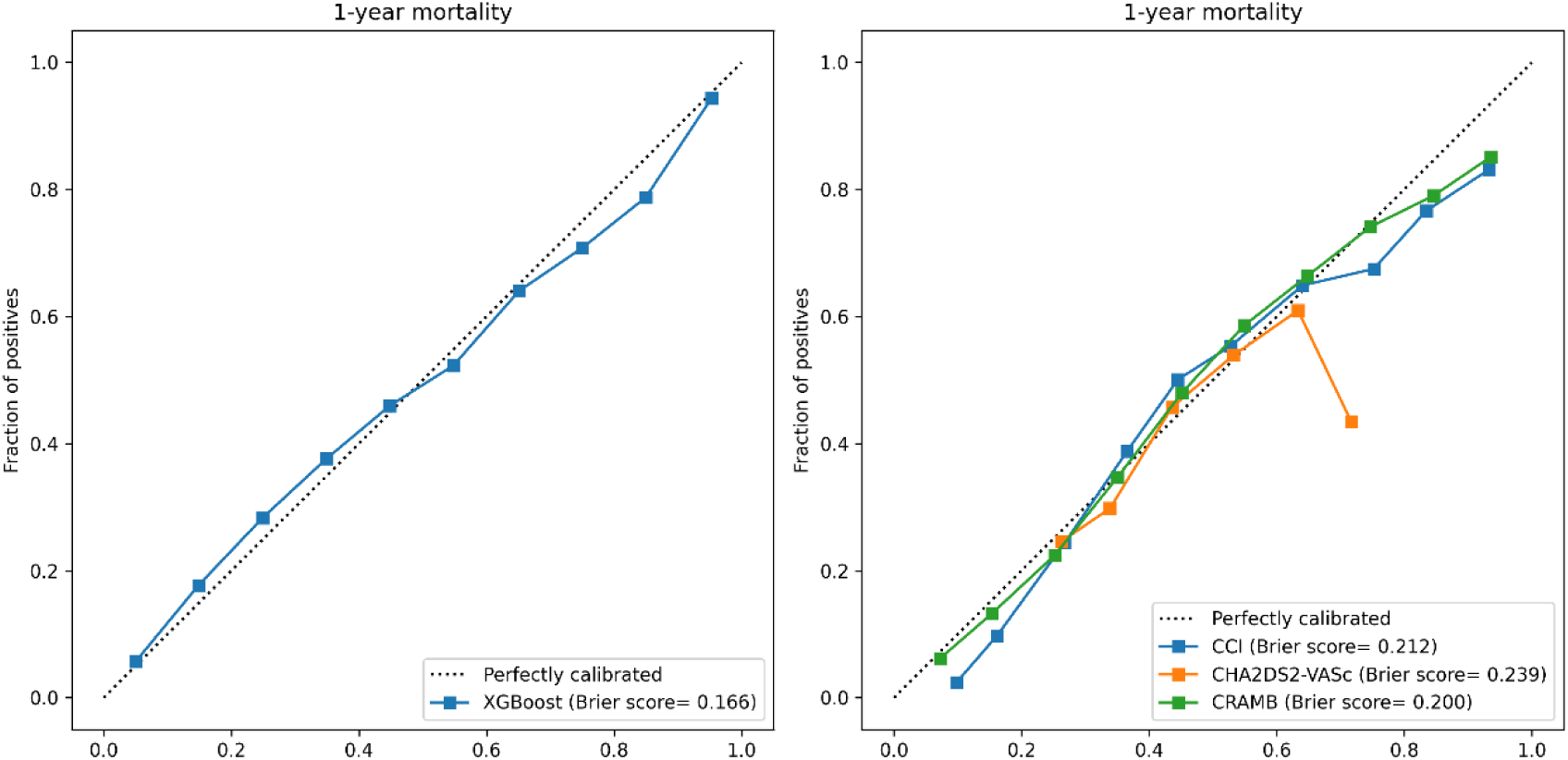
Calibration plots for the XGBoost model in the test set and the scoring model in the test set.

**Table 2.** Predictive performance of scores in the test set and DeLong test comparison. The p value for the Delong test was obtained by omparing the area under the curve (AUC) of the CRAMB score with of the corresponding score.

| **Item** | **Accuracy** | **Sensitivity** | **Specificity** | **ROC AUC** | **DeLong test P value** |
| --- | --- | --- | --- | --- | --- |
| **Charlson Comorbidity Index** | 0.659 | 0.563 | 0.738 | 0.720 | <0.001 |
| **CHA2DS2-VASc** | 0.582 | 0.397 | 0.733 | 0.609 | <0.001 |
| **CRAMB** | 0.690 | 0.594 | 0.768 | 0.756 | - |

**CHA2DS2-VASc score:** congestive heart failure, hypertension, age, diabetes mellitus, prior stroke or TIA or thromboembolism, vascular disease, age, sex category

### Discussion Main findings

This study’s primary contribution is the establishment of a benchmark for utilizing machine learning models in the construction of AF scores for mortality prediction. This study introduces and validates a novel risk score for assessing the 1-year mortality risk in patients with AF. By leveraging a large-sample population dataset and employing XGBoost models for predictor screening, the CRAMB (Charlson comorbidity index, readmission, age, metastatic solid tumor, and blood urea nitrogen maximum) score was developed. XGBoost excels in variable selection by effectively capturing nonlinear relationships and handling missing data [16]. Its built-in feature importance mechanism automatically identifies key variables, a capability lacking in logistic regression. Furthermore, compared with logistic regression, XGBoost’s ensemble learning often results in superior predictive performance, and its regularization techniques boost resilience against overfitting, making it a robust choice for predictive modeling and variable selection. The variables incorporated in the CRAMB score were validated through logistic and Cox regression analyses, demonstrating their predictive significance for mortality. The CRAMB score exhibited excellent calibration, and DCA illustrated its clinical utility. Importantly, the findings of this study revealed that the CRAMB score outperformed the widely used CHA2DS2-VASc risk score in predicting mortality, despite the latter’s original focus on predicting ischemic stroke.

### Predictors of death in patients with AF

Predictors and risk factors for death in patients with AF span a broad spectrum of clinical and demographic variables. Hypertension has been identified as a significant risk factor for incident heart failure and all-cause mortality in AF patients [17]. Moreover, an independent association has been observed between red cell distribution width and the risk of all-cause mortality in AF patients, with elevated red cell distribution width linked to increased mortality risk [18].

Left ventricular hypertrophy has been confirmed to be an independent risk factor for stroke and death in AF patients [19]. Moreover, patients with chronic kidney disease who develop AF face an increased risk of stroke and death [20], and renal function has been associated with the risk of stroke and bleeding in AF patients [21]. Additionally, age correlates with elevated risks of stroke and mortality in patients with either AF or sinus rhythm [22]. Those undergoing hemodialysis with new-onset AF exhibit heightened risks of death and stroke [23], and chronic kidney disease and hemodialysis impact mortality, length of stay, and total cost of hospitalization in patients admitted with a primary diagnosis of paroxysmal AF [24]. Furthermore, the biatrial volume predicts AF recurrence after ablation, impacting mortality [25].

Although a growing body of evidence indicates a connection between cancer and AF [26–28], the exact extent and underlying mechanisms of this association remain unclear. Proposed factors such as cancer-related inflammation, anticancer treatments, and other comorbidities associated with cancer are believed to influence atrial remodeling, potentially heightening the susceptibility of cancer patients to AF [26]. Specifically, new-onset AF has been associated with higher mortality in cancer patients [29, 30], and patients with solid tumors face an elevated risk of developing AF compared to those without cancer. The highest risk of AF occurs within 90 days of cancer diagnosis (OR 7.62, 95% CI 3.08 to 18.88), and this risk gradually decreases over time [30].

There is considerable evidence affirming the significance of BUN as a mortality predictor in patients with CVD [31–33]. Moreover, research indicates that BUN levels at admission serve as a predictive marker for mortality in patients with heart failure [32]. Elevated BUN levels (>13.51 mg/dL) in healthy older women have also been linked to a greater incidence of heart failure [34]. The present study contributes novel insights into the role of BUN in predicting mortality among AF patients through the use of the CRAMB score.

In summary, a review of the literature on predictor variables associated with AF death revealed that the variables used in constructing the CRAMB score are reasonable.

### Comparison with similar studies

Hijazi et al. proposed an innovative biomarker-based tool designed to predict mortality in patients with AF [8]. This score, named ABC-death (age, biomarkers (N-terminal pro B-type natriuretic peptide, troponin T, growth differentiation factor-15), and clinical history of heart failure), was developed and internally validated in 14,611 AF patients randomized to either apixaban or warfarin over a median period of 1.9 years. External validation was conducted in a cohort of 8,548 AF patients randomized to dabigatran or warfarin for 2.0 years. The study utilized a two-step variable selection method, initially fitting a model with all candidate predictors and then approximating a blinded, smaller model with the most predictive variables through ordinary least squares (OLS) and backward elimination. However, this approach has limitations, potentially excluding relevant variables and assuming linearity. Due to the exclusion of more severely ill patients in these two clinical trials, the scores developed from these datasets may not be applicable to other populations of severely ill patients with a greater number of comorbidities.

In a separate study, Samaras et al. introduced the BASIC-AF risk score (biomarkers, age, ultrasound, ventricular conduction delay, and clinical history), a novel prognostic tool for predicting mortality in AF patients [9]. This score was developed and validated using data from 1,130 patients in a single-center clinical trial; notably, the dataset was not split into training and test sets due to sample size restrictions. The study [9] incorporated traditional cardiac biomarkers and introduced echocardiography variables, identifying indexed left atrial volume as a significant predictor. This study [9] employed a random forest model for variable selection, and the random forest model has challenges with missing values, as it constructs decision trees based on specific features, posing difficulties in managing missing data. Imputing or estimating missing values introduces potential biases, and within the ensemble structure of random forests, this problem is compounded, compromising the model’s reliability in the case of incomplete data. XGBoost outperforms random forest models in variable selection because of its advanced regularization technique, gradient boosting framework for error correction, and more sophisticated tree-growing strategy. This approach results in improved predictive accuracy and precise estimation of variable importance.

Yan et al. focused on the development and validation of a prediction model for in-hospital death in patients with heart failure and AF [33]. Although that study did not directly align with the task of developing a score for risk of death in AF, it is relevant because it addresses the intersection of heart failure and AF, which are often comorbid conditions. The prediction model developed by Yan et al. may offer insights into prognostic factors and risk assessment strategies that may be applicable to the development of AF risk scores. The present study [33] developed a scoring model using 5998 patients admitted to the ICU from the MIMIC-IV database and used logistic regression to screen approximately 38 prespecified variables, ultimately including 16 variables to construct the score. However, the complexity of the scoring system prevented an evaluation of the importance of variables in variable screening based on traditional logistic regression.

Compared to the ABC-death risk score and BASIC-AF risk score, the CRAMB score was constructed based on the MIMIC-IV database, leading to significant differences in population characteristics compared to clinical trial populations. Directly comparing the AUC performance of the three scores is not appropriate. Although this study identified important predictor variables, such as the cardiac biomarker troponin T and comorbidities such as diabetes, their addition to the CRAMB score yielded only a marginal AUC benefit of less than 0.01 in absolute terms. Conversely, replacing or eliminating existing variables in the CRAMB score resulted in varying degrees of AUC performance reduction. This discrepancy may be attributed to the wide heterogeneity of the AF population in the MIMIC-IV database. Therefore, this study addresses a gap in the development of scoring methods and screening predictor variables within a broader population than did previous studies of this nature. Future research on AF scores should focus on the characteristics of the population used for score development, comprehensively considering the importance and applicability of the variables included.

### Limitations

The primary limitation of this study is its reliance on single-center data, preventing external validation of the developed AF score. Importantly, the predictor variables used for developing the AF risk score in this study may not be comprehensive enough. Additionally, the MIMIC-IV dataset includes derived databases such as MIMIC-IV-ECHO [35], which contains echocardiogram data; MIMIC-IV-ECG, which contains electrocardiogram data [36]; and MIMIC-IV-Note, which contains deidentified free-text clinical notes [37]. These derived datasets offer more patient characteristic data, enhancing the screening of variables.

### Conclusions

This study’s primary contribution is the establishment of a benchmark for utilizing machine learning models in the construction of a score for mortality prediction in AF patients. By leveraging a large-sample population dataset and employing XGBoost models for predictor screening, the CRAMB (CCI, readmission, age, metastatic solid tumor, and blood urea nitrogen maximum) score was developed. The CRAMB score’s simplicity makes it user-friendly, allowing for coverage of a broad and heterogeneous AF population. Moreover, the proposed model has superior predictive performance compared to that of the clinically utilized CHA2DS2-VASc risk score for 1-year mortality among AF patients. External validation of the CRAMB score in new datasets holds potential value for enhancing clinical practice.

## Data Availability

The data used in this study can be accessed on PhysioNet's website at https://physionet.org/content/mimiciv/2.2/.

## Acknowledgments

Not applicable.

## List of abbreviations

ABC-death: Age, biomarkers (N-terminal pro B-type natriuretic peptide, troponin T, growth differentiation factor-15), and clinical history of heart failure
AF: Atrial fibrillation
AUC: Area under the curve
BASIC-AF risk score: Biomarkers, age, ultrasound, intraventricular conduction delay, and clinical history
BUN: blood urea nitrogen
CCI: Charlson Comorbidity Index
CHA2DS2-VASc: congestive heart failure, hypertension, age, diabetes mellitus, prior stroke or TIA or thromboembolism, vascular disease, age, sex category
CI: Confidence interval
CRAMB: Charlson comorbidity index, readmission, age, metastatic solid tumor, and maximum blood urea nitrogen
DCA: Decision curve analysis
HR: hazard ratio
ICU: Intensive care unit
MIMIC-IV: Medical Information Mart for Intensive Care-IV
OR: Odds ratio
ROC: Receiver operating characteristic

